# Satisfaction levels with antenatal care services among postnatal women attending public hospitals in Sana’a City, Yemen

**DOI:** 10.64898/2026.09.04.26362239

**Authors:** Aisha Faisal Ahmed Al-Wadhaf, Nabil Ahmed Ahmed Al-Rabeei, Ghaida’a Ibrahim Ahmed Hafed, Lina Saeed Saif Al-Sofiani, Elham Haider Yusef Al-Jadi

## Abstract

**Background:** Women’s satisfaction with antenatal care (ANC) is an important indicator of maternal healthcare quality and is associated with the utilization of maternal health services. Identifying factors associated with satisfaction can help inform improvements in ANC service delivery. The study aims to assess women’s satisfaction and examine factors associated with ANC satisfaction among postnatal women attending public hospitals in Sana’a City, Yemen.

**Methods:** A descriptive cross-sectional study was conducted among 232 postnatal women attending three public hospitals in Sana’a City, Yemen. Participants were proportionally allocated across the three hospitals according to delivery volume, and eligible postnatal women were recruited consecutively until the required sample size was achieved. Data were collected through face-to-face interviews using a structured questionnaire. Descriptive statistics summarized participants’ characteristics and satisfaction levels. Chi-square tests assessed associations between categorical variables, and variables with p<0.05 in bivariate analysis were included in multivariable regression. Adjusted Odds Ratio (AORs) with 95% confidence intervals (CIs) were calculated, with statistical significance set at p<0.05.

**Results:** Overall, 105 of 232 women (45.3%) were classified as highly satisfied and 127 (54.7%) as having low satisfaction based on the median split. Several factors were independently associated with women’s satisfaction. Attendance at ANC education sessions was significantly associated with higher satisfaction, with women who attended sessions more likely to report satisfaction (AOR=6.60, p<0.001). Women with four or fewer children had higher odds of satisfaction (AOR=3.07, p=0.026). Waiting times of less than 30 minutes were associated with higher odds of satisfaction (AOR=3.14, p=0.024), while receiving a delivery plan was also positively associated with satisfaction (AOR=2.77, p=0.009). Residence and education were not significantly associated with women’s satisfaction after adjustment.

**Conclusions:** Women’s satisfaction with ANC was mainly associated with service-related and obstetric factors. Strengthening ANC education, reducing waiting times, and improving individualized care planning may improve women’s satisfaction with ANC services.

## Introduction

Women’s health is a fundamental component of public health because is directly associated with the wellbeing of women, newborns, families, and communities. Maternal health remains a major global health priority and is strongly emphasized within international development agendas. According to the World Health Organization (WHO), an estimated 260,000 women died worldwide from pregnancy- and childbirth-related causes in 2023 [1].

ANC refers to the health services provided by qualified health professionals to pregnant women and adolescent girls throughout pregnancy to promote the wellbeing of both the mother and the unborn child. These services include early detection of potential pregnancy risks, preventive, diagnostic, and therapeutic management of pregnancy-related conditions, as well as the provision of health education and counseling [2]. Effective ANC plays a vital role in reducing prenatal complications and encourages the utilization of skilled delivery and postnatal care services, which ultimately improves maternal and newborn health outcomes [3,4]. WHO recommends initiating ANC within the first trimester and attending at least eight visits during an uncomplicated pregnancy to optimize maternal and neonatal outcomes [2,5]. Timely and appropriate utilization of ANC services is important for improving maternal healthcare, and access to ANC varies according to individual and contextual factors [6]. Women’s satisfaction is an important indicator of the quality of ANC services because it reflects women’s experiences and perceptions of the care received [7]. Women’s satisfaction is widely recognized as an important indicator of healthcare quality because it reflects not only the technical competence of healthcare providers but also interpersonal communication, emotional support, privacy, respect, and responsiveness to women’s needs [2]. Higher levels of satisfaction have been associated with improved adherence to medical advice, greater utilization of maternal healthcare services, and better maternal and neonatal health outcomes [8].

However, despite global efforts to improve maternal healthcare, many low-resource and conflict-affected countries, including Yemen, continue to face major challenges related to accessibility, availability, and quality of ANC services. In Yemen, prolonged conflict, economic instability, and weaknesses in the healthcare system have negatively affected maternal healthcare services, including ANC. Limited healthcare resources, shortages of trained healthcare providers, long waiting times, and inadequate counseling services may be negatively associated with women’s experiences and satisfaction with ANC services. Assessing women’s satisfaction is therefore important for identifying gaps in service delivery and informing interventions aimed at improving the quality of maternal healthcare.

Although several international studies have examined women’s satisfaction with ANC services, evidence from Yemen remains limited. To the best of our knowledge, this is the first study conducted in Sana’a City, Yemen, to assess women’s satisfaction with ANC services and examine factors associated with satisfaction among postnatal women attending public hospitals. This study aimed to assess women’s satisfaction levels and examine factors associated with ANC satisfaction among postnatal women attending public hospitals in Sana’a City, Yemen.

## Materials and Methods

### Study design and setting

A descriptive cross-sectional study was conducted from January to March 2025 in three public hospitals in Sana’a City: Al-Thowrah Hospital, Al-Jomhury Hospital, and Al-Kuwait Hospital. One additional public hospital declined participation.

### Study population and sample size

The study population comprised all postnatal women admitted to the postnatal ward of target public hospitals in Sana’a City, Yemen, who delivered during the data collection period from January to March 2025. These women received maternal services during their hospital stay and were eligible for inclusion in the study. Sample size was calculated using Epi Info (StatCalc), population size (the total number of deliveries recorded across the three selected hospitals between October and December 2024) was 3,697; assuming an expected satisfaction level of 80% [8], a 95% confidence level, and a 5% margin of error, resulting in a required sample of 232 women.

### Sampling technique

A stratified proportional allocation approach followed by consecutive recruitment was used. The study population was stratified according to the participating hospitals, and the required sample size was proportionally allocated based on the number of deliveries in each hospital.

The total number of deliveries recorded across the three selected hospitals between October and December 2024 was 3,697. The sample size for each hospital was calculated using proportional allocation according to the following formula: *population size of the stratum ÷ total population size × total sample size*. This approach ensured proportional representation of women from each participating hospital.

Following institutional approval, eligible postnatal women were recruited consecutively in each hospital until the allocated sample size was reached. Consecutive recruitment was used because no complete sampling frame was available during the study period.

### Operational definitions

- Postnatal woman: Refers to a mother within the first six weeks after delivery.
- Women’s satisfaction: refers to a woman’s evaluation of the outcome resulting from a particular healthcare service and the care providers collectively.
- Level of women’s satisfaction: Refers to a measure of how healthcare services provided are perceived to meet or surpass women expectations.
- Healthcare providers (HCPs): Refer to physician/midwife or nurse who have been professionally certified in the skills required to manage deliveries and diagnose, manage or refer obstetric complications.
- ANC: Refers to care provided to pregnant women by a healthcare provider during pregnancy to promote and monitor the health and well-being of the mother and fetus.
- ANC follow-up: Refers to self-reported attendance at ANC during the current pregnancy (Yes/No).
- Attendance at ANC education sessions: Refers to self-reported attendance (Yes/No) at one or more structured health education sessions provided by healthcare providers during pregnancy. These sessions covered topics such as pregnancy care, nutrition, birth preparedness, delivery planning, and newborn care.
- Provision of a delivery plan: Refers to whether the ANC provider discussed and/or prepared an individualized delivery plan with the pregnant woman, including the intended place of delivery, skilled birth attendant, necessary supplies, and actions to be taken in the event of labor, complications, or other emergencies.

### Data collection

Data were collected through face-to-face interviews using a structured questionnaire adapted from previously validated instruments and relevant literature [9–15]. The questionnaire consisted of three main sections: socio-demographic characteristics, obstetric history, and women’s satisfaction with ANC services. The socio-demographic section included variables such as age, residence, educational status, occupation, and monthly family income in Yemeni Rials (YRs). Educational level was later recoded for logistic regression as uneducated (illiterate and able to read/write only) and educated (basic education, secondary education, diploma, and university education). The obstetric section, self-reported by women, included parity, number of children, pregnancy status, waiting time, ANC follow-up, attendance at ANC education sessions and provision of a delivery plan.

The variables “attendance at ANC education sessions” and “provision of a delivery plan” were collected as independent variables. Neither variable was included in the 18-item satisfaction scale used to calculate the overall satisfaction score. However, the satisfaction scale included an item assessing whether HCPs discussed skilled birth attendance and delivery planning. This item assessed women’s perceptions of communication and counseling during ANC and was distinct from the independent predictor “provision of a delivery plan,” which was based on whether the woman reported receiving an individualized delivery plan.

Women’s satisfaction with ANC services was assessed using 18 items grouped into nine dimensions: institutional structure, availability of HCPs, emotional support, cognitive support, promptness of care, confidentiality and privacy, interpersonal aspects of care, continuity of care, and cost of ANC services. The cognitive support dimension included discussion of regular ANC checkups, delivery planning, counseling on pregnancy danger signs, blood donor arrangements, adequacy of information provided, and HCPs willingness to answer questions.

To ensure data quality, field supervisors checked the completeness and accuracy of the questionnaires daily. The questionnaire was initially developed in English, translated into Arabic, and then back-translated into English to ensure consistency and linguistic accuracy. The tool was reviewed by academic experts in public health and obstetrics and gynecology. A pilot test (10%) was conducted prior to data collection to evaluate clarity and applicability of the instrument. The reliability of the satisfaction scale was assessed using Cronbach’s alpha coefficient. The obtained Cronbach’s alpha value was 0.909, indicating excellent internal consistency and reliability of the instrument.

### Data analysis

Data were entered and analyzed using IBM SPSS version 27. Descriptive statistics, including frequencies, percentages, means, and standard deviations, were used to summarize participants’ characteristics and levels of satisfaction with ANC services. Associations between women’s satisfaction and independent variables were initially examined using chi-square tests. When more than 20% of the expected cell counts were less than five, or when any expected cell count was less than one, likelihood-ratio chi-square or continuity-corrected chi-square tests were applied as appropriate. Effect sizes were evaluated using Phi coefficients Phi for 2x2 tables or Creamer’s *V* for 3x2 tables and interpreted according to Cohen’s criteria as weak (0.10–0.29), moderate (0.30–0.49), or strong (0.50–1.0).

Variables significant in the bivariate analysis (p<0.05) were entered into the multivariable logistic regression model using the enter method to identify factors independently associated with women’s satisfaction. AORs with 95% CIs were calculated. Multicollinearity among independent variables was assessed using the variance inflation factor (VIF), with values greater than four considered indicative of potential multicollinearity. All included variables demonstrated acceptable VIF values (<4), indicating no significant multicollinearity. To avoid conceptual overlap between predictors and the outcome measure, attendance at ANC education sessions and provision of a delivery plan were analyzed as independent predictors and were excluded from the computation of the overall satisfaction score. Educational level was analyzed in dichotomized form in both the bivariate and adjusted analyses. Statistical significance was set at p < 0.05. Model fitness was tested for goodness of fit using Hosmer–Lemeshow goodness-of-fit test and Nagelkerke R^2^ values.

The adequacy of the sample size for multivariable logistic regression was assessed using the events-per-variable (EPV) criterion. Six predictor variables were included and there were 105 women with high satisfaction, corresponding to an EPV of 17.5. The value exceeded the commonly used minimum of 10 events per variable, indicating an adequate number of outcome events relative to the number of predictors. There were no missing data for the variables included in the analysis. Reliability analysis of the satisfaction scale was conducted using Cronbach’s alpha, with values of 0.70 or higher considered acceptable.

#### Scoring system and categorization

Women’s satisfaction with ANC services was measured using 18 items distributed across nine dimensions. Responses were rated on a binary scale ranging from 1 (dissatisfied) to 2 (satisfied). Total satisfaction scores were calculated by summing the scores for all items. The median total satisfaction score was 32 and was used as the cutoff point for categorization. Women with scores less than or equal to the median score were categorized as having low satisfaction, whereas those with scores above the median score were categorized as having high satisfaction.

### Ethical considerations

Ethical approval for the study was obtained from the Research Ethics Committee of the Faculty of Medicine and Health Sciences, Al-Razi University, Sana’a, Yemen. Permission to conduct the study was also obtained from the administrations of the three participating hospitals. Verbal informed consent was obtained from all participants before data collection. The study included one participant aged 16 years and two participants aged 17 years. For these participants, the Research Ethics Committee approved independent informed consent rather than requiring guardian consent, based on their status as married adolescents. The participants were provided with information about the study and were considered capable of understanding the study procedures, risks, benefits, and their right to voluntarily decide whether to participate. Consent was documented in the consent section of the questionnaire by recording whether the participant agreed or declined to participate, with the interviewer’s signature confirming completion of the consent process. Participants were informed about the purpose and procedures of the study, their right to decline participation or withdraw at any time without affecting the care they received, and the confidentiality of the information provided. Anonymity and privacy were maintained throughout the study.

#### Ethics approval and consent to participate

The study was conducted in accordance with the ethical principles of the Declaration of Helsinki. The study protocol was approved by the Research Ethics Committee of the Faculty of Medicine and Health Sciences, Al-Razi University, Sana’a, Yemen [Approval No: RU/12/FMHS/2024]. The objective of the study was clearly explained to all participants. Verbal informed consent was obtained from all participants prior to inclusion in the study.

Confidentiality of participants’ information was strictly maintained and used solely for research and community benefit. All procedures were carried out in accordance with relevant guidelines and regulations.

## Results

### Socio-demographic characteristics of postnatal women

Table 1 describes the socio-demographic characteristics of the postnatal women. Participants’ ages ranged from 16 to 44 years (Mean = 27.22, SD = 6.14), with nearly half of the women aged 16–25 years (47.4%). The majority of respondents resided in urban areas (66.4%), while one-third lived in rural areas (33.6%).

**Table 1.**
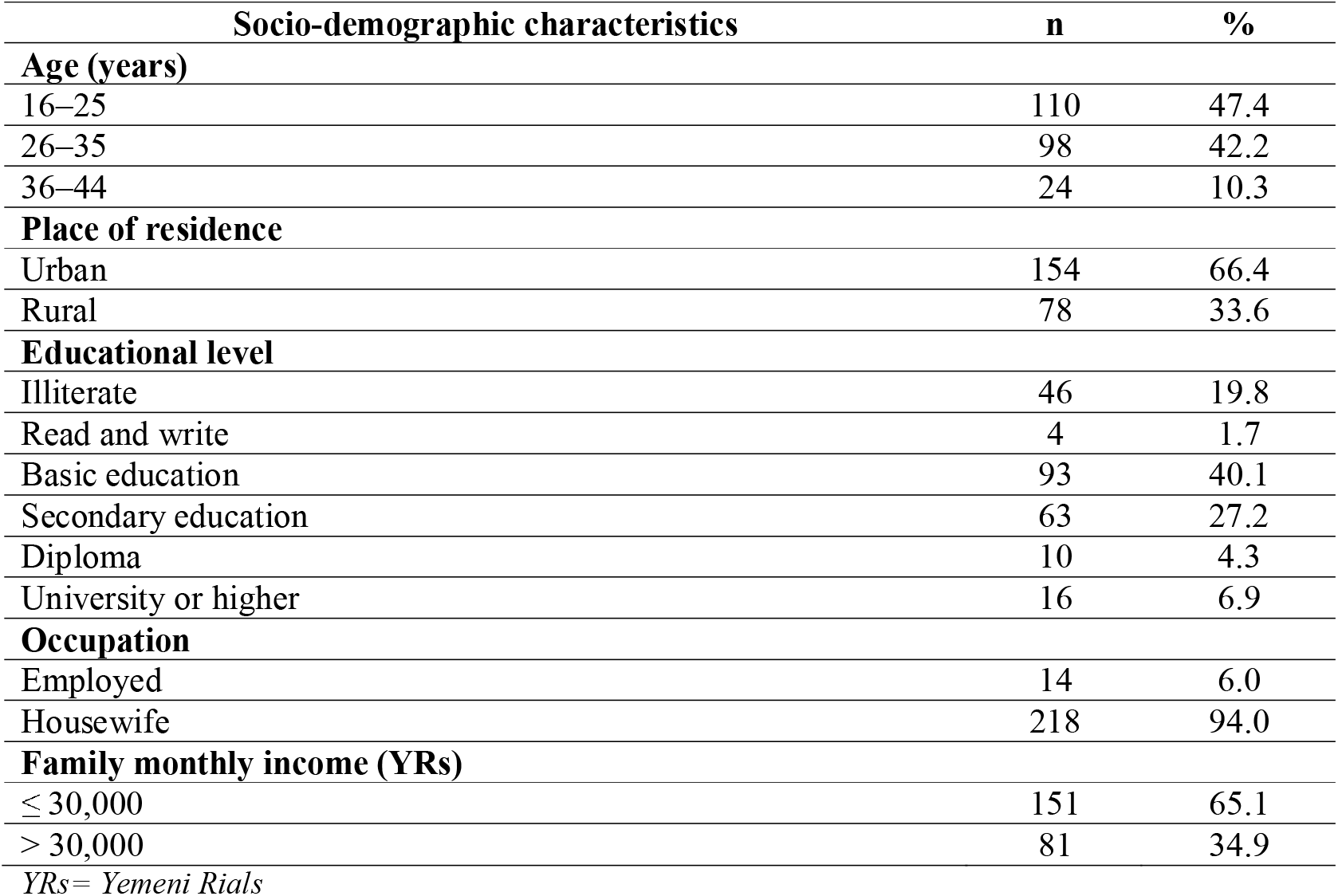
Socio-demographic characteristics of postnatal women.

| Socio-demographic characteristics | n | % |
| --- | --- | --- |
| **Age (years)** |  |  |
| 16–25 | 110 | 47.4 |
| 26–35 | 98 | 42.2 |
| 36–44 | 24 | 10.3 |
| **Place of residence** |  |  |
| Urban | 154 | 66.4 |
| Rural | 78 | 33.6 |
| **Educational level** |  |  |
| Illiterate | 46 | 19.8 |
| Read and write | 4 | 1.7 |
| Basic education | 93 | 40.1 |
| Secondary education | 63 | 27.2 |
| Diploma | 10 | 4.3 |
| University or higher | 16 | 6.9 |
| **Occupation** |  |  |
| Employed | 14 | 6.0 |
| Housewife | 218 | 94.0 |
| **Family monthly income (YRs)** |  |  |
| ≤ 30,000 | 151 | 65.1 |
| > 30,000 | 81 | 34.9 |

YRs= Yemeni Rials

Educational attainment was generally low, with the largest proportion of women having basic education (40.1%), followed by secondary education (27.2%). Approximately one-fifth of participants were illiterate (19.8%), whereas a small proportion had a university-level education or higher (6.9%). Most women were housewives (94.0%), with only 6.0% reporting formal employment. Regarding household income, nearly two-thirds of participants (65.1%) reported a monthly family income of 30,000 YRs or less.

### Obstetric characteristics of postnatal women

Table 2 presents the obstetric characteristics of the postnatal women included in the study. Nearly half of the participants were multiparous (46.6%), while 24.6% were primiparous and 28.9% were grand multiparous. The majority of women (81.5%) had four or fewer children. Most pregnancies were unplanned (80.2%). The majority of women experienced a waiting time of less than 30 minutes (84.5%). ANC follow-up was reported by most participants (93.1%); Attendance at ANC education sessions was reported by 51.7% of participants. However, only 32.3% reported having received a delivery plan during ANC, while more than two-thirds (67.7%) had not been provided with such a plan.

**Table 2.** Obstetric characteristics of postnatal women.

| Obstetric Characteristics | n | % |
| --- | --- | --- |
| **Parity** |  |  |
| Primipara | 57 | 24.6 |
| Multipara | 108 | 46.6 |
| Grand multipara | 67 | 28.9 |
| **Number of children** |  |  |
| ≤ 4 | 189 | 81.5 |
| > 4 | 43 | 18.5 |
| **Pregnancy status** |  |  |
| Planned | 46 | 19.8 |
| Unplanned | 186 | 80.2 |
| **Waiting time spent** |  |  |
| < 30 minutes | 196 | 84.5 |
| ≥ 30 minutes | 36 | 15.5 |
| **ANC follow-up** |  |  |
| Yes | 216 | 93.1 |
| No | 16 | 6.9 |
| **ANC education sessions** |  |  |
| Yes | 120 | 51.7 |
| No | 112 | 48.3 |
| **Provision of delivery plan** |  |  |
| Yes | 75 | 32.3 |
| No | 157 | 67.7 |

ANC = antenatal care

### Women’s satisfaction with ANC services

Table 3 summarizes women’s satisfaction levels across multiple dimensions of ANC services. Overall level of satisfaction, 105 of 232 women (45.3%) were classified as highly satisfied and 127 (54.7%) as having low satisfaction on the median split. More than four-fifths of participants were satisfied with the physical environment (81.9%) and cleanliness (81.5%) of the facilities. Regarding healthcare provider availability and emotional support, 75.4% of women were satisfied with the availability of healthcare providers, while 77.2% reported feeling comfortable with staff during ANC visits. Levels of satisfaction varied across the cognitive support dimension. Approximately two-thirds of women (68.1%) reported that healthcare providers discussed the importance of regular ANC checkups, and 70.7% indicated that providers were willing to answer questions. However, lower levels of satisfaction were reported regarding counseling on blood donor arrangements (32.3%), counseling on danger signs of pregnancy and childbirth (44.8%), and discussions related to skilled birth attendance and delivery planning (48.3%). Concerning promptness of care and privacy, 74.1% of women were satisfied with waiting times, and 74.6% were satisfied with the confidentiality and privacy maintained during ANC services. In terms of interpersonal aspects of care, most participants expressed satisfaction with the behavior and attitude of healthcare staff (76.3%) and reported feeling free to discuss their concerns during ANC visits (71.6%). Regarding continuity of care, 69.0% of women reported being encouraged to attend regular ANC follow-up visits, while 63.4% were satisfied with counseling related to nutrition and rest during pregnancy. Additionally, 65.9% felt adequately prepared for delivery. The lowest level of satisfaction was observed for the cost of ANC services, where only 44.0% of women reported satisfaction, while 56.0% expressed dissatisfaction.

**Table 3.** Satisfaction with ANC services among postnatal women attending public hospitals in Sana’a City, Yemen.

| Dimensions of satisfaction | Satisfied<br>n (%) | Dissatisfied<br>n (%) |
| --- | --- | --- |
| **Institutional structure** |  |  |
| Physical environment of the ANC facility | 190(81.9) | 42 (18.1) |
| Cleanliness of the ANC facility | 189(81.5) | 43 (18.5) |
| **Availability of HCPs** | 175(75.4) | 57 (24.6) |
| **Emotional support** |  |  |
| Felt comfortable with staff during ANC visits | 179(77.2) | 53 (22.8) |
| **Cognitive support** |  |  |
| HCPs discussed the importance of regular ANC checkups | 158(68.1) | 74 (31.9) |
| HCPs discussed skilled birth attendance and delivery planning | 112(48.3) | 120 (51.7) |
| HCPs provided counseling on danger signs of pregnancy and childbirth | 104(44.8) | 128 (55.2) |
| HCPs discussed blood donor arrangements | 75 (32.3) | 157 (67.7) |
| Adequacy of information provided during ANC | 128(55.2) | 104 (44.8) |
| HCPs were willing to answer questions | 164(70.7) | 68 (29.3) |
| **Promptness of care** |  |  |
| Satisfaction with waiting time | 172(74.1) | 60 (25.9) |
| **Confidentiality and privacy** | 173(74.6) | 59 (25.4) |
| **Interpersonal aspects of care** |  |  |
| Behavior and attitude of healthcare staff | 177(76.3) | 55 (23.7) |
| Felt free to discuss all concerns | 166(71.6) | 66 (28.4) |
| **Continuity of care** |  |  |
| Encouraged to attend regular ANC follow-up visits | 160(69.0) | 72 (31.0) |
| HCPs discussed nutrition and adequate rest during pregnancy | 147(63.4) | 85 (36.6) |
| Felt adequately prepared for delivery | 153(65.9) | 79 (34.1) |
| **Cost of ANC services** | 102(44.0) | 130 (56.0) |
| *HCPs= Healthcare providers; ANC = antenatal care* |  |  |

### Association between women’s satisfaction with ANC and characteristics

Table 4 presents the association between women’s satisfaction with ANC services and socio-demographic and obstetric characteristics. Place of residence was significantly associated with women’s satisfaction with ANC services (*p* = 0.020), with a weak effect size (φ = 0.15), indicating that women residing in urban areas were more likely to report higher satisfaction than those living in rural areas. Educational level also demonstrated a significant association with satisfaction (*p* = 0.033), with a weak effect size (φ = 0.14). This suggests that women’s satisfaction varied by educational attainment, although the strength of this relationship was modest.

**Table 4.**
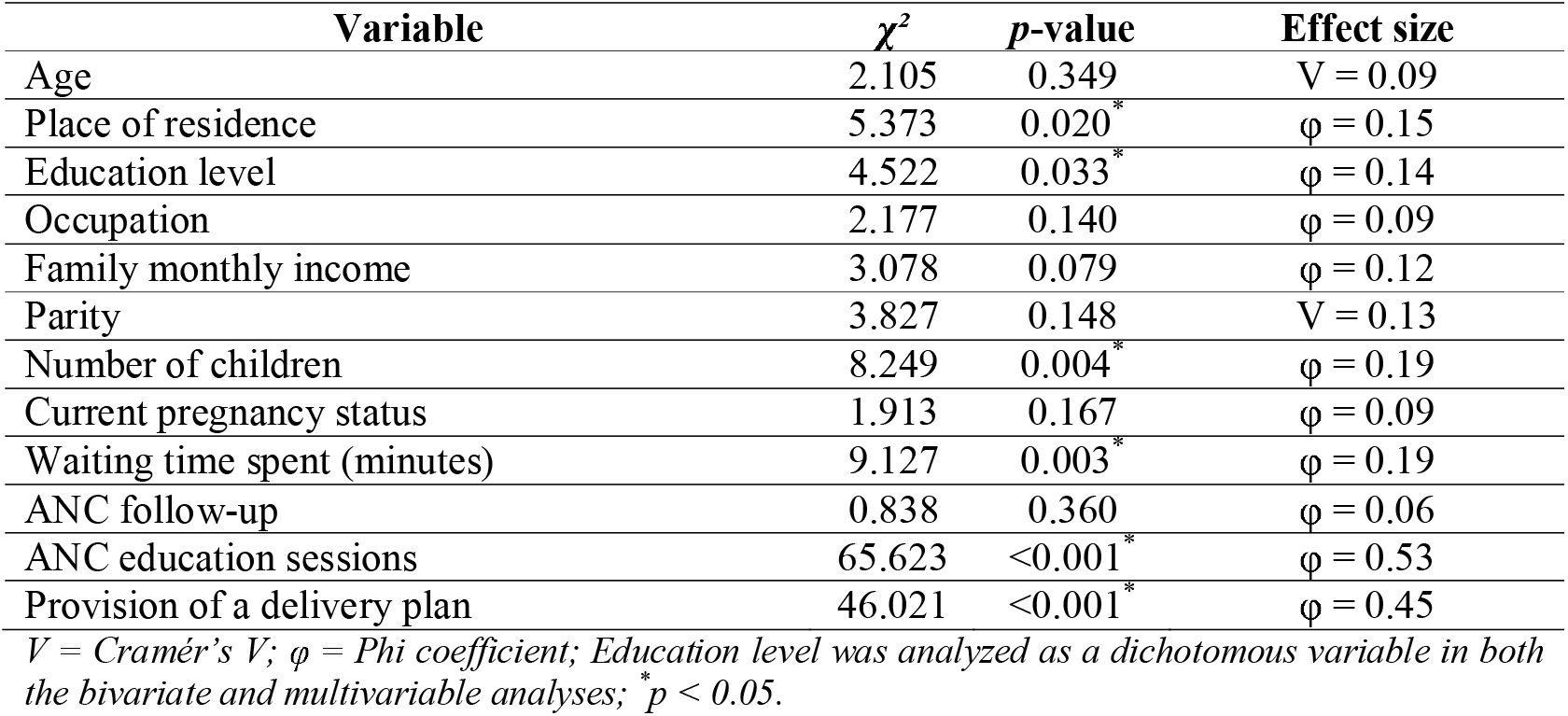
Association between women’s satisfaction with ANC and socio-demographic and obstetric characteristics.

A significant association was found between number of children and women’s satisfaction with ANC (*p*= 0.004), with a weak effect size (φ= 0.19). Women with fewer children reported higher satisfaction compared with those who had more children.

Waiting time was also significantly associated with satisfaction (*p* = 0.003), with a weak effect size (φ= 0.19). Shorter waiting times were associated with higher levels of satisfaction among women receiving ANC services. Attendance at ANC education sessions demonstrated a strong and statistically significant association with women’s satisfaction (*p* < 0.001), with a large effect size (φ = 0.53). Women who attended ANC education sessions were substantially more likely to report high satisfaction compared with those who did not attend such sessions. Similarly, provision of a delivery plan was significantly associated with women’s satisfaction (*p* < 0.001), with a moderate effect size (φ = 0.45). Women who received a delivery plan during ANC reported higher satisfaction levels than those who did not receive such planning. No significant associations were observed with age, occupation, parity, pregnancy status, or ANC follow-up (all p > 0.05).

### Multivariable logistic regression analysis

A multivariable logistic regression analysis was performed to identify factors associated with women’s satisfaction with ANC services among postnatal women attending public hospitals in Sana’a City, Yemen (Table 5). The overall model was statistically significant (Omnibus test: χ^2^ = 88.572, p< 0.001). The model explained 42.4% of the variance in women’s satisfaction (Nagelkerke R^2^= 0.424), and demonstrated adequate fit (Hosmer–Lemeshow test: χ^2^= 6.226, p =0.514). The model correctly classified 76.3% of participants. After adjusting for potential confounders, several factors remained independently associated with women’s satisfaction with ANC services.

**Table 5.**
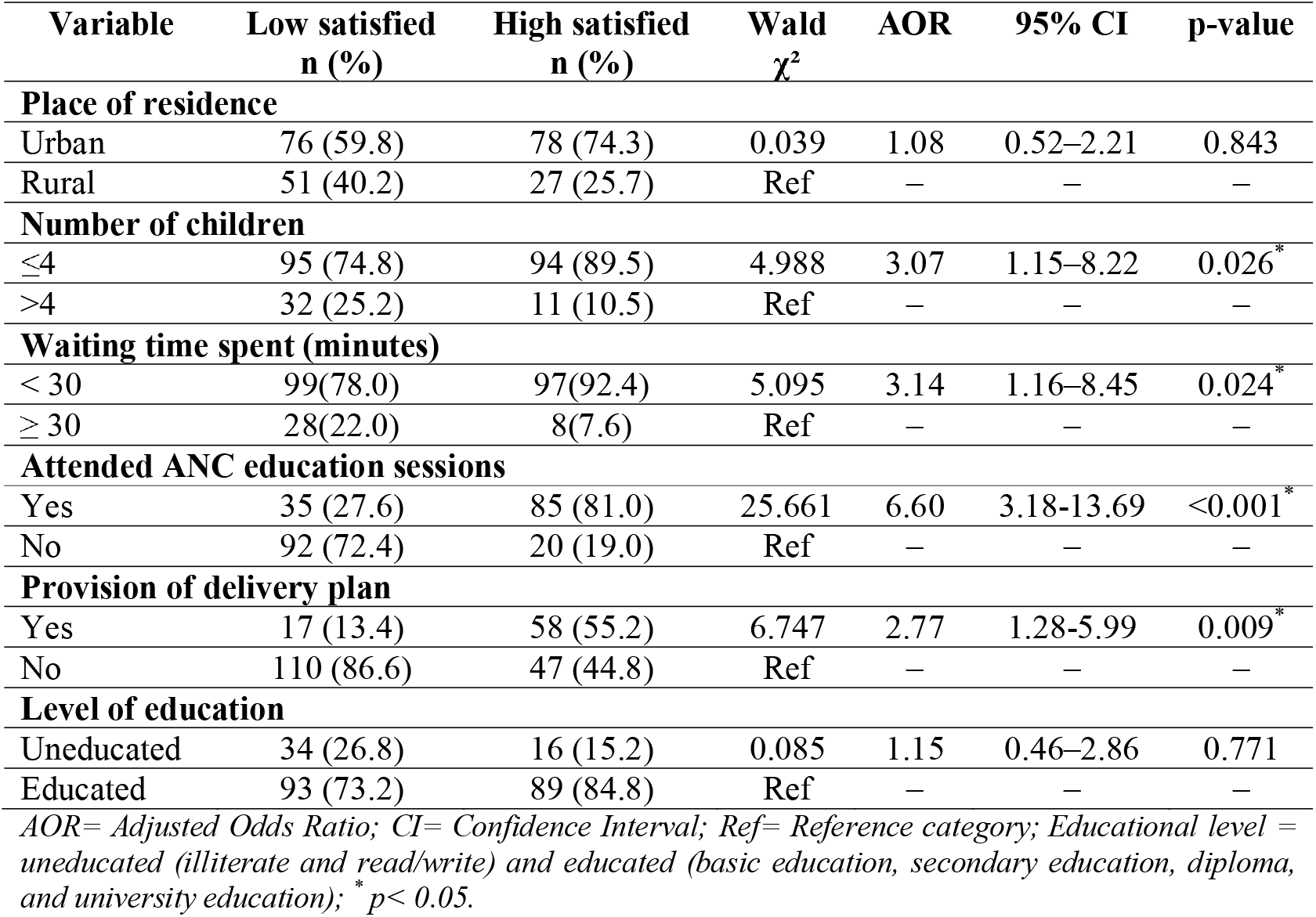
Factors associated with women’s satisfaction with ANC services among postnatal women attending public hospitals in Sana’a City, Yemen.

Attendance at ANC education sessions was independently associated with satisfaction. Women who attended ANC education sessions were significantly more likely to report high satisfaction than those who did not attend. Specifically, 81.0% of highly satisfied women had attended ANC education sessions compared with 27.6% among women with low satisfaction. Women who attended ANC education sessions had 6.60 times higher odds of being highly satisfied with ANC services compared with those who did not attend (AOR = 6.60, 95% CI: 3.18–13.69, p < 0.001). Number of children was also independently associated with satisfaction. Women with four or fewer children constituted 89.5% of the highly satisfied group compared with 74.8% of the low-satisfaction group. Participants with ≤ 4 children were approximately 3.07 times more likely to report high satisfaction compared with women who had more than four children (AOR = 3.07, 95% CI: 1.15–8.22, p = 0.026).

Waiting time was independently associated with women’s satisfaction with ANC services. Women who waited <30 minutes represented 92.4% of the highly satisfied group compared with 78.0% of the low-satisfaction group. Women who waited less than 30 minutes were 3.14 times more likely to report high satisfaction than those who waited 30 minutes or longer (AOR = 3.14, 95% CI: 1.16–8.45, p = 0.024). Provision of a delivery plan was another significant predictor of satisfaction. Women who received a delivery plan were more likely to report high satisfaction compared with those who did not receive one (AOR = 2.77, 95% CI: 1.28–5.99, p = 0.009). A delivery plan was reported by 55.2% of highly satisfied women compared with 13.4% of women with low satisfaction.

In contrast, place of residence and educational level were not significantly associated with satisfaction level after adjustment for other variables in the model. Although educational level was statistically significant in the bivariate analysis, this association was no longer significant after adjustment for other variables in the model, suggesting that its apparent association with satisfaction was explained by other covariates included in the multivariable model.

## Discussion

This study identified several factors associated with women’s satisfaction with ANC services in public hospitals in Sana’a City, Yemen. The findings showed that attendance at ANC education sessions, receiving a delivery plan, shorter waiting time, and lower number of children were significant predictors of higher woman’s satisfaction with ANC services.

Attendance at ANC education sessions was the strongest predictor of satisfaction. Women who participated in these sessions were more likely to report positive experiences with ANC services than those who did not attend. Health education may contribute to improved understanding of pregnancy-related issues, available health services, and self-care practices. It also enhances communication between women and healthcare providers, increases confidence in the care received, and encourages active participation in healthcare decisions. Similar findings have been reported in previous studies, which found that effective communication and educational support positively associated with women’s satisfaction [16,17].

Provision of a delivery plan was also significantly associated with greater satisfaction. Women who received individualized delivery planning reported better experiences with ANC services. Delivery plans may be associated with better preparedness for childbirth, increased sense of autonomy, and greater trust in healthcare providers. These findings suggest that women’s experiences with ANC are associated more strongly with the quality and responsiveness of services than with socioeconomic characteristics. Comparable findings were reported by Ayalew et al. [18] and Mutianingsih and Mardiningsih [19], who emphasized the importance of respectful maternity care and birth preparedness in improving maternal satisfaction.

The study further revealed that women with four or fewer children were more satisfied with ANC services compared with women who had more than four children. Women with fewer children may receive greater attention from healthcare providers and may be more engaged in ANC services during pregnancy. Similar findings were reported by Akinyemi et al. [20]. However, evidence regarding parity and maternal satisfaction remains inconsistent across studies, suggesting that social and cultural contexts may be associated with women’s perceptions of care.

Waiting time was another important determinant of satisfaction. Women who waited less than 30 minutes were more likely to report satisfaction than those who experienced longer waiting periods. Short waiting times may indicate better service organization, improved provider responsiveness, and higher perceived quality of care. This finding is consistent with studies conducted in other low-resource settings, where prolonged waiting times were linked to dissatisfaction with maternal healthcare services [21]. Similarly, Adedeji et al. [11] reported that health education sessions, provider attitude, waiting time, and cost of services were important determinants of ANC satisfaction.

Although residence was associated with satisfaction in the bivariate analysis, it did not remain significant after adjustment in the multivariable model. This suggests that differences between urban and rural women may largely be explained by variations in service quality and access rather than residence itself. Urban women reported higher satisfaction than rural women, reflecting better access to healthcare facilities and services in urban areas [6].

The study also found no significant association between educational level and woman’s satisfaction after controlling for confounding variables. While women with higher education may have greater expectations regarding healthcare services, the present findings indicate that interpersonal care and service quality may play a more important role in shaping satisfaction than educational background alone. Similarly, occupation, family income, pregnancy intention, and ANC follow-up visits were not significantly associated with woman’s satisfaction. These findings suggest that women’s experiences with ANC are associated with the quality and responsiveness of services than by socioeconomic characteristics alone. Comparable findings were reported by Kifle et al. [22] and Kassahun et al. [23].

Overall, the findings highlight the multidimensional nature of maternal satisfaction with ANC services. Improving satisfaction requires strengthening health education, reducing waiting times, promoting individualized delivery planning, and ensuring respectful and responsive maternity care. Such interventions may contribute to increased utilization of maternal healthcare services and improved maternal and neonatal outcomes in Yemen and similar low-resource settings.

### Strengths and limitations

This study provides important evidence on factors associated with women’s satisfaction with ANC services in Sana’a City, Yemen, where limited research has been conducted. The inclusion of socio-demographic, obstetric, and service-related variables enhanced the comprehensiveness of the analysis. In addition, the use of multivariable regression allowed adjustment for potential confounding factors and identification of independent predictors of woman’s satisfaction. However, some limitations should be considered. The cross-sectional design limits the ability to establish causal relationships between the identified factors and satisfaction with ANC services. The information obtained may have been impacted by recall bias due to the self-reported nature of the data. In addition, the study was conducted only in selected public hospitals in Sana’a City, which may limit the generalizability of the findings to private healthcare facilities or rural areas outside the study setting. Although attendance at ANC education sessions and provision of a delivery plan were analyzed as independent predictors and excluded from the satisfaction score, these variables are conceptually related to women’s perceptions of care. Therefore, some overlap in interpretation cannot be completely excluded. Despite these limitations, the study offers useful evidence to support maternal healthcare planning and policy development in Yemen.

## Conclusions

Women’s satisfaction with ANC services in Sana’a City was independently associated with attendance at ANC education sessions, receiving a delivery plan, shorter waiting times, and lower number of children. Service-related factors appeared to show stronger associations with women’s satisfaction than socio-demographic characteristics. Strengthening health education during ANC visits, improving delivery planning, and reducing waiting times may enhance women’s experiences and improve the quality of maternal healthcare services. Policymakers and healthcare providers should adopt comprehensive woman-centered approaches to strengthen ANC services and improve maternal health outcomes in Yemen.

## Acknowledgement

The authors would like to express their sincere gratitude to all the mothers who participated in this study for their time and cooperation. We also thank the administrations and healthcare staff of Al-Thowrah Hospital, Al-Jomhury Hospital, and Al-Kuwait Hospital for granting permission and facilitating data collection.

## Data availability statement

The de-identified dataset underlying the findings of this study has been deposited in Zenodo and is publicly available at https://doi.org/10.5281/zenodo.21376026

## Consent for publication

Not applicable. This study does not contain any individual person’s data in any form (including individual details, images, or videos).

## Abbreviations

ANC: Antenatal Care
HCPs: Healthcare Providers
VIF: Variance Inflation Factor
AORs: Adjusted Odds Ratio
CIs: Confidence Intervals
WHO: World Health Organization
YRs: Yemeni Rials
Φ: Phi coefficient
V: Creamer’s V
EPV: Events-Per-Variable.

## Author contributions

Aisha Faisal Ahmed Al-Wadhaf, MSc. Degree in Public Health

**Roles:** First and corresponding author, Conceptualization, Investigation, Methodology, Project administration, Validation, Writing review & editing, Writing original draft.

**Affiliation:** Al-Razi University, Faculty of Medicine and Health Sciences, Community Health and Nutrition Department

Nabil Ahmed Ahmed Al-Rabeei, Professor in Public Health and Dietetics

**Roles:** Supervision, Formal analysis, Methodology, Investigation, Software, Visualization, Writing original draft

**Affiliation:** Al-Razi University, Faculty of Medicine and Health Sciences, Community Health and Nutrition Department Ghaida’a Ibrahim Ahmed Hafed, MSc. Degree in Public Health

**Roles:** Data collection, Data entry, Validation

**Affiliation:** Al-Razi University, Faculty of Medicine and Health Sciences, Community Health and Nutrition Department Lina Saeed Saif Al-Sofiani, MSc. Degree in Epidemiology

**Roles:** Data collection, Data entry, Validation

**Affiliation:** Al-Razi University, Faculty of Medicine and Health Sciences, Community Health and Nutrition Department Elham Haider Yusef Al-Jadi, MSc. Degree in Public Health

**Roles:** Data collection, Data entry, Validation

**Affiliation:** Al-Razi University, Faculty of Medicine and Health Sciences, Community Health and Nutrition Department

## References

1. World Health Organization. Maternal mortality. Geneva: World Health Organization; 2025. Available from: https://www.who.int/news-room/fact-sheets/detail/maternal-mortality Accessed 1 July 2026.

2. World Health Organization. WHO recommendations on antenatal care for a positive pregnancy experience. Geneva: World Health Organization; 2016. Available from: https://www.who.int/publications/i/item/9789241549912. Accessed July 2, 2026.

3. Kassa ZY, Chojenta C, Loxton D, Tegegne TK. Women’s satisfaction with antenatal care in sub-Saharan Africa: A systematic review and meta-analysis. BMC Pregnancy Childbirth. 2020;20:702. doi:10.1186/s12884-020-03369-1.

4. Bintabara D, Nakamura K, Seino K, Kizuki M. Determinants of institutional delivery and antenatal care services utilization in Tanzania: a community-based cross-sectional study. Int J Womens Health. 2019;11:353–365. doi:10.2147/IJWH.S207699.

5. Odusina EK, Ahinkorah BO, Ameyaw EK, Seidu AA, Budu E, Zegeye B, et al. Noncompliance with the WHO’s recommended eight antenatal care visits among pregnant women in sub-Saharan Africa: a multilevel analysis. Biomed Res Int. 2021;2021:6696829. doi:10.1155/2021/6696829. PMID:34589549.

6. Awoke N, Ababulgu SA, Hanfore LK, Gebeyehu EG, Wake SK. Regional disparities in antenatal care utilization among pregnant women and its determinants in Ethiopia. Front Glob Womens Health. 2024;5:1230975. doi:10.3389/fgwh.2024.1230975.

7. Lakew S, Ankala A, Jemal F. Determinants of client satisfaction to skilled antenatal care services at Southwest of Ethiopia: a cross-sectional facility-based survey. BMC Pregnancy Childbirth. 2018;18:479. doi:10.1186/s12884-018-2121-6.

8. Addisalem K, Tamirat G, Firehiwot M, Addis E, Merga D. Maternal satisfaction on delivery care services and associated factors at public hospitals in eastern Ethiopia. Int Health. 2023;15(2):189–197. doi:10.1093/inthealth/ihac038.

9. Alemu EM, Kaso AW, Obsie GW, Fessaha HZ, Agero G. Maternal satisfaction with delivery service and associated factors among women who gave birth at public hospitals in Guji Zone, Southern Ethiopia. BMC Womens Health. 2024;24:227. doi:10.1186/s12905-024-03069-0.

10. Hosseinzadeh M, Pouladzadeh M, Eskandari A. Assessment of healthcare service quality and patient satisfaction using the SERVQUAL questionnaire in Khuzestan Province during 2022–2023. Jundishapur J Chronic Dis Care. 2024;13(4):e146329. doi:10.5812/jjcdc-146329.

11. Adedeji OA, Oluwasola TAO, Adedeji FM. Assessment of antenatal care satisfaction amongst postpartum women at the University College Hospital, Ibadan, Nigeria. Eur J Obstet Gynecol Reprod Biol X. 2023;20:100252. doi:10.1016/j.eurox.2023.100252.

12. Okari MG. Satisfaction levels with maternity services among postnatal women attending public hospitals in Nairobi City County, Kenya [master’s thesis]. Nairobi: Kenyatta University; 2018.

13. USAID. Women’s level of satisfaction with maternal health services in Jharkhand, India: Findings from the quantitative study. Task Order No. GHS 1 01 07 00016 00. New Delhi: USAID; 2012.

14. Navas A, Blanco-López L, Seoane-Pillado T, López-Castiñeira N, Díaz SP. Women’s satisfaction with childbirth and postpartum care and associated variables. Rev Esc Enferm USP. 2021;55:e03720. doi:10.1590/S1980-220X2020006603720.

15. Panagopoulou V, Kalokairinou A, Tzavella F, Tziaferi S. A survey of Greek women’s satisfaction of postnatal care. AIMS Public Health. 2018;5(2):158–172. doi:10.3934/publichealth.2018.2.158.

16. Rahman MM, Ngadan DP, Arif MT. Factors affecting satisfaction on antenatal care services in Sarawak, Malaysia: Evidence from a cross-sectional study. SpringerPlus. 2016;5:725. doi:10.1186/s40064-016-2437-3.

17. Panth A, Kafle P. Maternal satisfaction on delivery service among postnatal mothers in a government hospital, Mid-Western Nepal. Obstet Gynecol Int. 2018;2018:4530161. doi:10.1155/2018/4530161.

18. Ayalew MM, Nebeb GT, Bizuneh MM, Dagne AH. Women’s satisfaction and its associated factors with antenatal care services at public health facilities: a cross-sectional study. Int J Womens Health. 2021;13:279–286. doi:10.2147/IJWH.S293725.

19. Mutianingsih R, Mardiningsih S. The characteristics level of maternal satisfaction in receiving antenatal care. Int J Midwifery Res. 2022;2(2). doi:10.47710/ijmr.v2i2.32.

20. Akinyemi JO, Afolabi RF, Awolude OA. Patterns and determinants of dropout from maternity care continuum in Nigeria. BMC Pregnancy Childbirth. 2016;16:282. doi:10.1186/s12884-016-1083-9.

21. Kebede DB, Belachew YB, Selbana DW, Gizaw AB. Maternal satisfaction with antenatal care and associated factors among pregnant women in Hossana Town. Int J Reprod Med. 2020;2156347. doi:10.1155/2020/2156347.

22. Kifle MM, Ghirmai FA, Berhe SA, Tesfay WS, Weldegebriel YT, Gebrehiwet ZT. Predictors of women’s satisfaction with hospital-based intrapartum care in Asmara public hospitals, Eritrea. Obstet Gynecol Int. 2018; 3717408. doi:10.1155/2018/3717408.

23. Kassahun A, Berhe A, Gebremedhin S. The effect of antenatal care on use of institutional delivery service and postnatal care in Ethiopia: a systematic review and meta-analysis. BMC Health Serv Res. 2018; 18:577. doi:10.1186/s12913-018-3370-9.

